# Diagnostic Role of Wall Thickness Heterogeneity in Hypertrophic Cardiomyopathy and in Carriers of Sarcomeric Gene Mutations

**DOI:** 10.1101/2025.07.31.25332464

**Authors:** Giovanni Donato Aquaro, Roberto Licordari, Giancarlo Todiere, Umberto Ianni, Carmelo De Gori, Marco Merlo, Andrea Barison, Antonio De Luca, Crysanthos Grigoratos, Francesca Raimondi, Lamia Ait-Ali, Pierluigi Festa, Francesco Negri, Calogero Falletta, Francesco Bianco, Fabrizio Ricci, Nicoletta Botto, Lapo Taddei, Mattia Alberti, Andrea Italiano, Luna Gargani, Francesco Clemenza, Lorenzo Faggioni, Dania Cioni, Riccardo Lencioni, Michele Emdin, Emanuele Neri, Gianfranco Sinagra, Raffaele De Caterina, Gianluca Di Bella

## Abstract

**Background:** Hypertrophic cardiomyopathy (HCM) is characterized by variable left ventricular hypertrophy, but current diagnostic criteria rely on absolute wall thickness thresholds that may miss early or subtle phenotypic expressions, especially in sarcomere mutation carriers.

**Objectives:** This study aimed to assess whether wall thickness standard deviation (WTSD), as a marker of wall thickness heterogeneity, improves the identification of HCM and mutation carriers.

**Methods:** We evaluated 382 healthy controls, 297 patients with guideline-based HCM diagnosis, and 82 sarcomere mutation carriers without overt hypertrophy using cardiac magnetic resonance imaging. End-diastolic wall thickness (WT) was measured across 16 myocardial segments, and WTSD was calculated. Diagnostic performance of WTSD and other wall thickness-derived parameters was assessed using age- and sex-specific thresholds.

**Results:** WTSD was significantly higher in HCM patients (4.3 ± 1.1 mm) and mutation carriers (2.3 ± 0.3 mm) compared to healthy controls (1.3 ± 0.3 mm; p<0.0001). WTSD identified 97% of HCM patients and 64% of mutation carriers, with excellent specificity (99%). In females, WTSD achieved a sensitivity of 98.9% and specificity of 100% for HCM diagnosis, and identified 74% of female mutation carriers. WTSD outperformed demographic-based personalized thresholds and BSA-indexed maximal WT in all subgroups. Mutation carriers exhibited increased heterogeneity due to the coexistence of hypertrophic and thinned segments, despite normal absolute WT.

**Conclusions:** WTSD is a robust imaging biomarker that detects early and overt manifestations of sarcomeric HCM with high accuracy. Incorporating WT heterogeneity into diagnostic algorithms may enhance early identification, particularly in women and mutation carriers.

**CONDENSED ABSTRACT:** Wall thickness heterogeneity, quantified as wall thickness standard deviation (WTSD) by magnetic resonance, emerged as a powerful diagnostic marker of sarcomeric hypertrophic cardiomyopathy (HCM). In 297 HCM patients and 82 mutation carriers, WTSD identified nearly all overt cases and 64% of carriers without hypertrophy with excellent specificity (99%). Particularly in women, WTSD achieved near-perfect diagnostic accuracy and uncovered early disease otherwise missed by conventional thresholds. These findings support incorporating WT heterogeneity into diagnostic algorithms to enable earlier identification of at-risk individuals and refine risk stratification beyond absolute wall thickness criteria.

## INTRODUCTION

Hypertrophic cardiomyopathy (HCM) is the most prevalent inherited cardiac disease and leading cause of sudden cardiac death (SCD) in younger person (1).

HCM is a primary cardiomyopathy characterized by increased ventricular wall thickness unexplained by underlying systemic, metabolic and cardiac conditions (such as hypertension and aortic stenosis) (2). Although HCM is most often an inherited monogenic cardiac disease that is transmitted as an autosomal dominant trait, incomplete penetrance and variable expressivity are common with many individuals experiencing minimal symptoms, while others developing heart failure, atrial fibrillation or SCD (3). In adults, HCM is diagnosed in the setting of an end diastolic (ED) maximal wall thickness (WT) ≥15 or ≥13 mm in presence of family history or pathogen mutation (2).

These thickness cut-off values do not take into account several factors. The majority of HCM cases present with asymmetric hypertrophy, with 16 different patterns described (4), and there are known differences in the distribution of normal values based on ethnicity, sex, and body size (e.g., women and individuals of Asian descent tend to have relatively smaller hearts) (5,6). There is growing evidence that sex contributes to explain some of the clinical heterogeneity seen among HCM patient. Indeed, females present at a more advanced stage of disease, have higher symptom burden, carry greater risk for heart failure and, most importantly, for mortality compared to males (7,8), probably because the thresholds for wall thickness are too high in females and only identify patients in more advanced stages of the disease. In a recent study, Shiwani and colleagues proposed an approach to adjust the threshold of ED maximal WT based on demographic characteristics such as sex, age, and body surface area (BSA) (9). Other studies have proposed using wall thickness indexed to body surface area (10,11).

Another phenotypic characteristic of HCM is the heterogeneity in wall thickness, where some segments appear hypertrophic while others are thinned. This aspect has been poorly investigated. We recently performed post-mortem cardiac magnetic resonance (CMR) on explanted hearts from patients with HCM who died from sudden cardiac death (12). In these hearts, due to *rigor mortis*, the left ventricle is captured at a variable phase of diastole depending on the ATP reserves at the time of death, making it impossible to measure ED maximal WT . In that study, we showed that the standard deviation (SD) of wall thickness across all left ventricular segments was significantly higher in HCM patients compared to healthy controls and patients with hypertrophy from other causes (12).

The aim of the present study is: a) to identify the normal range of the SD of left ventricular end-diastolic wall thickness (WTSD) in a population of healthy subjects; and b) to assess whether WTSD, as a marker of wall thickness heterogeneity, may have a role in the diagnosis of HCM by evaluating a cohort of patients with a definite diagnosis of HCM and a group of patients with a pathogenic mutation in sarcomeric genes but without a diagnosis of HCM (mutation carriers).

## Methods

### Study population: healthy controls

A population of 400 healthy controls were enrolled using the following inclusion criteria: age ≥ 18 years, no family history of cardiac disease, normal electrocardiogram, no history of cardiac diseases or symptoms, no cardiovascular risk factors (hypertension, dyslipidemia, diabetes, obesity, smoking, family history), no known systemic diseases, and no contraindications to CMR. Healthy controls were divided by sex and by age class (<35 years, ≥35 to <50 years, ≥50 to <65 years, ≥65 years). Four patients excluded from the analysis for suboptimal image quality and 14 for the finding of minor abnormal tissue or mophological features at CMR.

### Study population: HCM/mutation carriers

A population of 300 patients aged ≥18 years with a guideline-based diagnosis of HCM was enrolled: patients with diagnosis of HCM defined as having ED maximal WT ≥15 mm in any myocardial segment that is not explained solely by loading conditions (with or without known pathogen mutation of sarcomere gene) or with diagnosis of HCM determined by ED maximal WT of ≥13 mm and <15 mm with a pathogen mutation of sarcomere gene (2).

A group of 82 “mutation carriers” aged ≥18 years with ED maximal WT <13 mm but with a known pathogen mutation of sarcomere gene and family history of HCM and with the following exclusion criteria: no personal history of other cardiac diseases or cardiovascular symptoms, no cardiovascular risk factors (hypertension, dyslipidemia, diabetes, obesity, smoking, family history), no known systemic diseases, and no contraindications to CMR.

Overall, 3 patients with guideline-base diagnosis of HCM were excluded from the analysis suboptimal image quality (2 frequent premature ventricular complexes during the acquisition of cine-SSFP images, 1 for atrial fibrillation during CMR).

The study was approved by the internal ethical committee, and all subjects gave their written informed consent. All the patients enrolled underwent clinical, electrocardiographic, and echocardiographic evaluation at the time of CMR.

### CMR acquisition protocol

CMR scan was performed using different 1.5 Tesla machines with dedicated cardiac coil. Study protocol included functional evaluation with short axis cine images, acquired from the mitral plane valve to the LV apex, and these images were acquired using a steady-state free precession pulse sequence with the following parameters: 30 phases, slice thickness 8 mm, no gap, views per segment 8, FOV 35-40 cm, phase FOV 1, matrix 224×224, reconstruction matrix 256×256, 45° flip angle, and a TR/TE near to 2. Cine images with the same parameters were acquired also in 2-, 3- and 4-chambers views. Late Gadolinium enhancement (LGE) images were acquired about 10 minutes after the administration of 0.5 molar gadolinium contrast agent (0.2 mmol/kg) in short and long-axis views, using an inversion recovery T1-weighted gradient-echo sequence or a Phase Sensitive Inversion recovery pulse sequence, acquired with the following parameters: field of view 35 to 40 mm, slice thickness 8 mm, no gap, repetition time 3 to 5 ms, echo time 1 to 3, a flip angle of 20°, matrix 224×224, and reconstruction matrix 256×256. Using a TI-scout, the appropriate inversion time identify to null normal myocardium.

### Post-processing image analysis

All CMR studies were analyzed offline by 3 experienced CMR readers with level III EACVI accreditation for CMR, blinded to all clinical data. A commercially available research software package (CVI42, Circle Cardiovascular Imaging Inc., Calgary, Canada) was used to quantify functional parameters. LV mass was measured by the analysis of short-axis cine images by manually tracing endocardial and epicardial contours of LV myocardium in the end-diastolic and end-systolic phases. End-diastolic volume index end-systolic volume index, mass, and mass index were measured as previously described. LV end-diastolic WT of each of the 16 conventional AHA/ACC/ESC myocardial segment was automatically measured by using the endocardial and epicardial contours of short axis slices after removing the papillary muscle from LV mass (13). Intracavitary trabeculation was included in LV cavity and only “compacted” myocardium was included in LV walls. Briefly, in each end-diastolic short axis slice 100 chordae were automatically traced orthogonally to myocardial wall from the endocardial to the epicardial contour (**figure 1**). The ED maximal WT, the minimal WT, the mean WT and the standard deviation (WTSD) were obtained for each myocardial segment from the analysis of these chordae as respectively, the maximal, the minimal, the mean and the SD of the chordae. The ED maximal WT was also indexed for body surface area (maximal WT-index). For each patient the “demographic-based personalized threshold” of ED maximal WT was considered based on the body surface area, the class of age and sex, considering both the 95% and 99.7% PI of normal range, as recently proposed by Shiwani and coworker (9).

**Figure 1:**
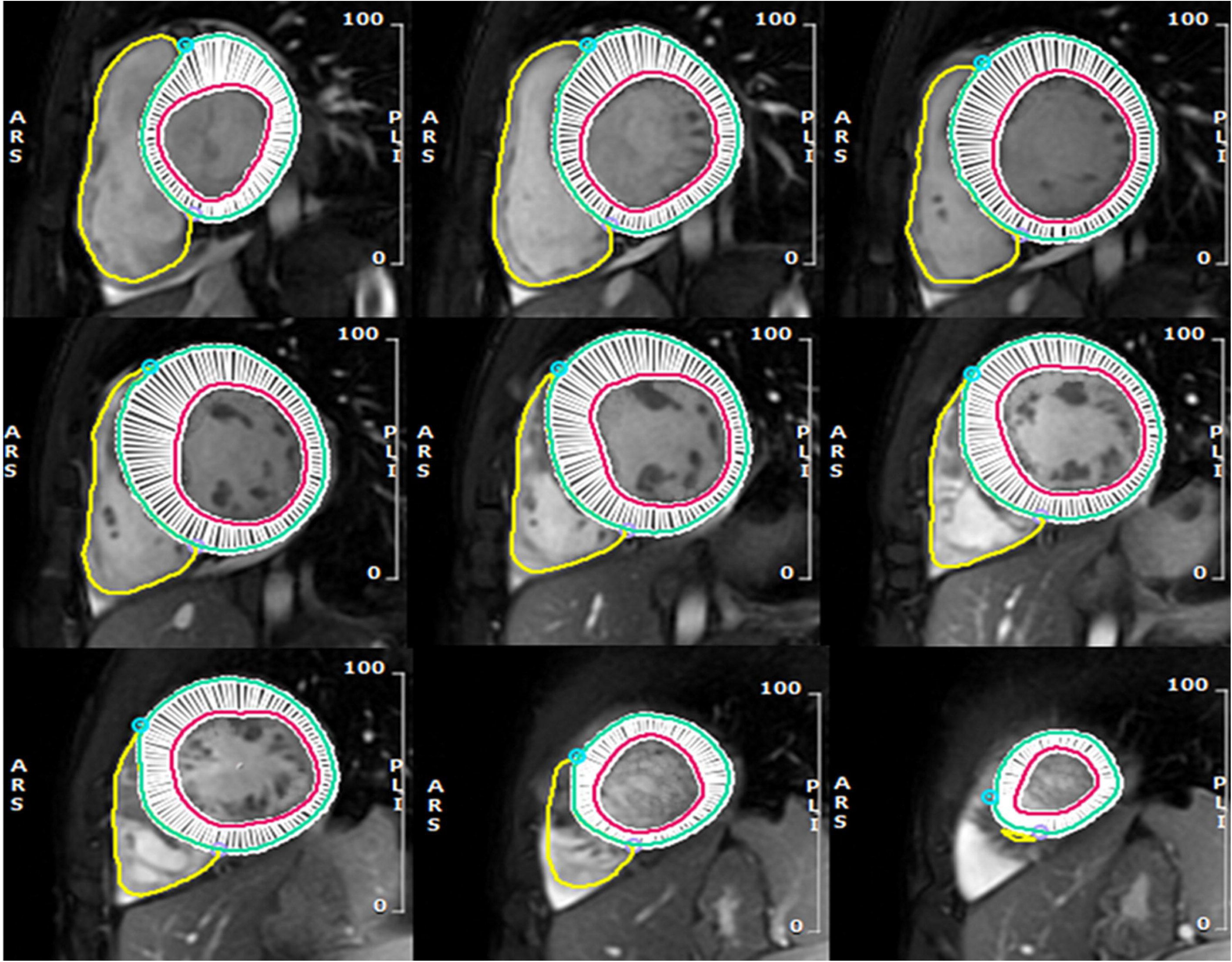
Example of cardiac magnetic resonance image analysis. Endocardial and epicardial contours were traced on short-axis ventricular images at end-diastole. The software then automatically generated 100 radial chords per slice to measure wall thickness. For each myocardial segment, the maximum and minimum end-diastolic wall thickness were derived from these chords. Then the mean end-diastolic wall thickness and the standard deviation (SD) of segmental wall thickness were calculated.

For apical hypertrophy, the ratio apical\basal end-diastolic thickness was measured for each of the 6 LV walls (anterior, inferior, anterolateral, inferolateral, anteroseptal, inferoseptal). Particularly, for the anterolateral and inferolateral it was measured as ratio of distal lateral segment and, respectively, the basal anterolateral and inferolateral wall, and for the anteroseptal and inferoseptal wall as the ratio of the distal septum and, respectively, the basal anteroseptal and inferoseptal wall. For each patient, the maximal apical\basal ratio measured was considered in the analysis as continuous variables and using 3 differerent thresholds: >1, ≥1.3 and ≥1.5. Finally, the ED maximal thickness of the four apical segments was indexed to BSA, and the reference cut-off values recently proposed by Hughes and coworkers for healthy subjects were applied: ≤5.2 mm for the apical anterior and lateral segments, 5.3 mm for the apical inferior segment, and 5.6 mm for the apical septum (10.).

### Statistical analysis

Values are presented as mean ± SD or as median and 25^th^-75^th^ for variables with normal and non-normal distribution, respectively. A Kolmogorov-Smirnov test for normal distribution was used.

All the parameters derived from ED WT measured in healthy controls, with the exception of the maximal-minimalWT difference, had a normal distribution in each one of the four classes of age and in both sexes. Based on these findings, thresholds of normality for each parameter were calculated as mean plus 2 SD. The 95° percentile was, instead, used for the maximal-minimal WT difference as threshold.

The chi-square test or the Fisher exact test were used, when appropriate, for categorical variables. Comparison among groups for continuous variables were made with the Student’s t test or 1-way analysis of variance or the Kruskal-Wallis test as appropriate.

Sensitivity, specificity, positive predictive and negative predictive values, area under the curve (AUC) and the diagnostic accuracy was measured for all the wall thickness derived parameters to identify HCM patients or mutation carriers. AUC comparison was used to compare effectiveness of each parameters to identify HCM or mutation carriers.

## RESULTS

### Population

The final study population included 382 healthy controls (216 males, 51±15 years old), 297 patients with guideline-based diagnosis of HCM and 82 carriers of sarcomere pathogen mutations with a family history of HCM.

Characteristics of the population of healthy controls, HCM patients and mutation carriers are reported in Table **1**. As evident in that table, among the patients with definite diagnosis of HCM 226 out of 297 had a sarcomere-positive mutation: 164 a pathogenic variant and 62 a pathogenic- like variant.

**Table 1:** Characteristics of the study populations.

| Variables |  | Healthy controls | HCM | Carriers of pathogenic sarcomeric mutation | Healthy. vs HCM P value | Healthy. vs Mutation carriers P value |
| --- | --- | --- | --- | --- | --- | --- |
| n. |  | 382 | 297 | 82 |  |  |
| Age (years) | mean ± SD | 49(34-61) | 52(32-69) | 45(28-56) | 0.24 | 0.03 |
| Males | n (%) | 216 (56%) | 205(51%) | 28(34%) | 0.41 | <0.0001 |
| Systemic Hypertension | n (%) | 0 (0%) | 5 | 0 | 0.02 | - |
| - | n (%) | 0 (0%) | 1 | 0 | 0.29 | - |
| Diabetes | n (%) | 0 (0%) | 1 | 0 | 0.29 | - |
| Smoking | n (%) | 0 (0%) | 19 | 5(6%) | 0.29 | - |
| Sarcomere-positive pathogenic variants | n (%) | 0(0%) | 164(55%) | 82(100%) | <0.0001 | <0.0001 |
| Sarcomere-positive like-pathogenic variants | n (%) | 0(0%) | 62(21%) | 0(0%) | <0.0001 | - |
| Genetic analysis not performed | n (%) | 350(100%) | 71(24%) | 0(0%) | <0.0001 | <0.0001 |
| Family History of HCM | n(%) | 0 | 156(53%) | 82(100%) | <0.0001 |  |
| ED maximal WT (mm) | mean ± SD | 9.8 ± 1.6 | 18±3 | 10.2±1.6 | <0.0001 | 0.04 |
| ED maximal WT < 13 mm | n (%) | 382 (100%) | 0 | 82 (100%) | <0.0001 | - |
| ED maximal WT ≥ 13 & <15 mm | n (%) | 0 | 41(14%) | 0 | <0.0001 | - |
| ED maximal WT ≥ 15 mm | n (%) | 0 | 256(86%) | 0 | <0.0001 | - |
| ED mean WT (mm) | mean ± SD | 7.1 ± 1.2 | 8.2±2 | 5.7± 1.2 | <0.0001 | 0.001 |
| ED minimal WT(mm) | mean ± SD | 4.9 ± 1.3 | 4.9±1.4 | 3.8±1 | 0.85 | <0.0001 |
| ED maximal-minimal WT (mm) | median (IQR) | 4.8(4-6) | 13(10-15) | 6.5(5.6-7.6) | <0.0001 | <0.0001 |
| WTSD | | 1.3±0.3 | 4.3±1.1 | 2.3±0.3 | <0.0001 | <0.0001 |
| Maximal WT indexed by BSA | mm/m<sup>2</sup> | 5.1±1 | 9.8±2 | 6.0±1.1 | <0.0001 | <0.0001 |
| LV mass index | g/m<sup>2</sup> | 69±13 | 93±18 | 64± 11 | <0.0001 | 0.17 |
| LV EDVi (ml) | mL/m<sup>2</sup> | 75±12 | 68±11 | 71± 16 | 0.002 | 0.25 |
| LV ESVi (ml) | mL/m<sup>2</sup> | 25(19-32) | 21(15-29) | 21(19-32) | 0.0007 | 0.009 |
| LV EF | % | 66(62-71) | 72(63-76) | 70(62-71) | 0.004 | 0.03 |
| RV EDVi | mL/m<sup>2</sup> | 76(65-91) | 66(58-77) | 67(65-71) | <0.0001 | 0.03 |
| RV EF | % | 63(58-68) | 64(58-71) | 65(64-75) | 0.14 | 0.08 |
| LV atrium (cm2) | cm<sup>2</sup> | 11(10-13) | 13(11-16) | 11(10-13) | <0.0001 | 0.6 |
| LVoutflow obstruction | n (%) | 0 | 37(12) | 0 | <0.0001 | - |
| Septal hypertrophy | n (%) | 0 | 220(74%) | 0 | <0.0001 | - |
| Apical hypertrophy | n (%) | 0 | 45(15%) | 0 | <0.0001 | - |
| Diffuse hypertrophy | n (%) | 0 | 29(10%) | 0 | <0.0001 | - |
| Inferior/inferolateral hypertrophy | n (%) | 0 | 3(1%) | 0 | <0.0001 | - |
| Late Gadolinium Enhancement | n (%) | 0(0%) | 212(71%) | 0 | <0.0001 | - |

HCM, Hypertrophic cardiomyopathy; ED, end-diastolic; WT, wall thickness; WTSD, standard deviation of the wall thickness; BSA, body surface area; LV, left ventricle; EDVi, End-diastolic volume index; ESVi, End-systolic volume index; EF, ejection fraction

Healthy controls had an obvious lower ED maximal WT than HCM patients but, interestingly, also lower than mutation carriers. Data from Table 1 suggest that both HCM patients and mutation carriers are characterized by greater heterogeneity in WT, as the WTSD was significantly higher in both groups, 4.3±1.1 and 2.3±0.3 respectively, compared to healthy controls (WTSD 1.3±0.3, p<0.0001 for both comparisons). Additional evidence of this heterogeneity is provided by the significantly greater difference maximal-minimal WT observed in HCM patients (median 13, range 10–15) and mutation carriers (median 6.5, range 5.6–7.6) compared to healthy controls (median 4.8, range 4–6, p<0.0001 in both comparisons). Interestingly, in mutation carriers, who, by definition, did not have myocardial segments with WT ≥ 13 mm, this heterogeneity is associated with the presence of segments with reduced wall thickness. Indeed, both the minimal WT (3.8±1 mm vs. 4.9±1.3 mm, p<0.0001) and the mean WT (5.7±1.2 mm vs. 7.1±1.2 mm, p=0.001) were significantly lower in mutation carriers compared to healthy controls. On contrast, in HCM mean WT was greater (p<0.0001) and the minimal WT was not significantly different (p=0.85) than healthy controls. Finally, the maximal WT indexed by BSA was greater in both HCM and mutation carriers than healthy controls (p<0.0001 in both cases). An example of CMR analysis in a healthy control, a patient with HCM, and a mutation carrier is shown in **Central Illustration**.

Parameters derived from ED WT measured in healthy controls are shown in table 2. The thresholds of normality of each parameter was obtained, as explained in statistical analysis section, from this population of healthy controls divided for sex and class of age and are reported in Table **3**.

**Table 2:** Wall thickness parameters in healthy controls divided by age class.

| Variable: | Age Class, years |  |  |  | P value |
| --- | --- | --- | --- | --- | --- |
|  | <35 | ≥35 & <50 | ≥50 & <65 | ≥65 |  |
| **<u>Males:</u>** |  |  |  |  |  |
| ED maximal WT | 8.2±1.7 | 9.3±1.1 | 9.3±1.2 | 9±1.4 | <0.0001 |
| ED minimal WT | 4.7±1.7 | 5.4±1.1 | 5.5±0.9 | 5.7±0.8 | <0.0001 |
| Maximal-minimal WT | 4.7(3.7-5.4) | 5(4.1-5.6) | 5.3(3.9-5.7) | 4.9(4.2-6.6) | 0.048 |
| ED mean WT | 6.9±1.5 | 7.8±1.0 | 7.7±1.1 | 7.5±1.2 | <0.0001 |
| ED WTSD | 1.18±0.23 | 1.31±0.25 | 1.51±0.39 | 1.51±0.4 | <0.0001 |
| ED maximal WT index* | 5.1±0.6 | 5.0±0.5 | 5.4±0.8 | 5.7±0.79 | <0.0001 |
| Max apex/base ratio | 0.66±0.17 | 0.68±0.12 | 0.63±0.16 | 0.73±0.11 | 0.08 |
| Apical max WT index | 3.5±0.6 | 3.5±0.7 | 3.6±0.8 | 3.7±1 | 0.12 |
| **<u>Females:</u>** |  |  |  |  |  |
| ED maximal WT | 7.9±0.8 | 8.8±1 | 9.2±0.8 | 9.1±0.9 | <0.0001 |
| ED minimal WT | 3.9±0.74 | 4.3±0.9 | 4.2±1.1 | 5.3±1.02 | <0.0001 |
| Maximal-minimal WT | 4.0(3.3-4.4) | 4.8(3.8-5.4) | 4.9(4.3-6.8) | 4.6(3.5-5.1) | <0.0001 |
| ED mean WT | 5.6±0.79 | 6.4±0.91 | 6.7±1.01 | 6.6±1.4 | 0.002 |
| ED WTSD | 0.99±0.18 | 1.19±0.21 | 1.33±0.27 | 1.29±0.27 | <0.0001 |
| ED maximal WT index* | 4.8±0.33 | 5.3±0.91 | 5.3±0.69 | 5.5±0.79 | <0.0001 |
| Max apex/base ratio | 0.63±0.1 | 0.66±0.12 | 0.62±0.12 | 0.76±0.11 | 0.08 |
| Apical max WT index | 3.2±0.7 | 3.3±0.9 | 3.5±0.8 | 3.8±0.9 | 0.18 |

ED, end-diastolic, WT wall thickness, SD standard deviation.

**Table 3:** Normality range of end-diastolic wall thickness parameters.

|  | Age Class ( years) |  |  |  |
| --- | --- | --- | --- | --- |
|  | <35 | ≥35 &<50 | ≥50 & <65 | ≥65 |
| **<u>MALES:</u>** |  |  |  |  |
| **ED maximal WT** | <12 | <12 | <12 | <12 |
| **ED minimal WT (mean +2SD)** | ≤8 | ≤8 | ≤7 | ≤7 |
| **Maximal-minimal WT (95°)** | ≤6 | ≤6 | ≤8 | ≤8 |
| **ED mean WT (mean +2SD)** | ≤10 | ≤10 | ≤10 | ≤10 |
| **ED WTSD (mean +2SD)** | ≤1.63 | ≤1.79 | ≤2.28 | ≤2.3 |
| **ED maximal WT index (mean +2SD)** | ≤6.3 | ≤6 | ≤7 | ≤7.3 |
| **Max apex/base ratio (mean +2SD)** | ≤1 | ≤0.92 | ≤0.95 | ≤0.95 |
| **Apical max WT index (mean +2SD)** | ≤4.7 | ≤4.9 | ≤5.2 | ≤5.7 |
| **<u>FEMALES:</u>** |  |  |  |  |
| **ED maximal WT** | <10 | <11 | <11 | <11 |
| **ED minimal WT (mean +2SD)** | ≤5 | ≤6 | ≤6 | ≤7 |
| **Maximal-minimal WT (95°)** | ≤6 | ≤6 | ≤7 | ≤7 |
| **ED mean WT (mean +2SD)** | ≤7 | ≤8 | ≤9 | ≤9 |
| **ED WTSD (mean +2SD)** | ≤1.36 | ≤1.62 | ≤1.86 | ≤1.83 |
| **ED maximal WT index (mean +2SD)** | ≤5.5 | ≤7.1 | ≤6.8 | ≤7.1 |
| **Max apex/base ratio (mean +2SD)** | ≤0.83 | ≤0.9 | ≤0.86 | ≤0.94 |
| **Apical max WT index (mean +2SD)** | ≤4.6 | ≤5.1 | ≤5.1 | ≤5.6 |

ED, end-diastolic, WT wall thickness, SD standard deviation.

*Wall-thickness derived parameters in the whole population of patients with definite HCM diagnosis* Table 4 shows the evaluation of the diagnostic performance of wall thickness-derived parameters in identifying HCM patients and carriers of pathogenic sarcomeric mutations compared to healthy controls, using the corresponding age- and sex-specific reference cut-offs provided in Table 3. The diagnostic performance of the recently proposed Demographic-Based Personalized Thresholds of maximal WT(cut-offs >95th and >99th percentile) (9) was also assessed. Sensitivity, specificity, AUC, PPV and NPV were calculated for each parameter.

**Table 4:** Diagnostic accuracy of wall thickness derived parameters in the whole population of HCM and mutation carriers.

| **Patients with guideline-based diagnosis of HCM vs controls:** |  |  |  |  |  |  |
| --- | --- | --- | --- | --- | --- | --- |
|  | **Sensitivity** | **Specificity** | **AUC** | **PPV** | **NPV** | **AUC**<br>**comparis**<br>**on vs**<br>**WTSD**<br>**(P value)** |
| Demographic-Based<br>Personalized Thresholds of<br>maximal WT (95%PI)* | 84(79-87) | 99.4(98-99.9) | 0.91(0.89-0.94) | 99.2(97-99.8) | 87(85-90) | <0.0001 |
| Demographic-Based<br>Personalized Thresholds of<br>maximal WT (99.7%PI)* | 73(67-78) | 99.7(98-99.9) | 0.86(0.83-0.89) | 99.5(97-99.9) | 82(80-85) | <0.0001 |
| WTSD | 97(94-98) | 99(97-99.7) | 0.98(0.96-0.99) | 98.6(96-99.6) | 97(95-99) | - |
| ED minimal WT | 6(4-10) | 97(94-98) | 0.51(0.47-0.55) | 59(42-74) | 56(50-57) | <0.0001 |
| Maximal-minimal WT | 88(84-91) | 90(86-92) | 0.89(0.86-0.91) | 89(86-92) | 88(85-91) | <0.0001 |
| ED mean WT | 41(36-47) | 97(94-98) | 0.69(0.66-0.73) | 91(85-95) | 67(66-70) | <0.0001 |
| ED maximal WT index | 97(95-99) | 89.5(86-92) | 0.93(0.91-0.95) | 87(84-91) | 97(95-98) | 0.0001 |
| Maximal apex/base ratio | 16(9-21) | 99(96-99.9) | 0.57(0.52-0.62) | 95(71-99) | 58(52-55) | <0.0001 |
| Apical maximal WT index | 15(11-21) | 99(98-99.9) | 0.59(0.53-0.61) | 96(84-99) | 63(59-61) | <0.0001 |
| **Carriers of sarcomere pathogen mutation without HCM diagnosis vs controls:** |  |  |  |  |  |  |
|  | **Sensitivity** | **Specificity** | **AUC** | **PPV** | **NPV** | **AUC**<br>**comparis**<br>**on vs**<br>**WTSD**<br>**(P value)** |
| Demographic-Based<br>Personalized Thresholds of<br>maximal WT (95%PI)* | 11(5-20) | 99.4(98-99.9) | 0.55(0.51-0.60) | 82(50-95) | 84(83-85) | <0.0001 |
| Demographic-Based<br>Personalized Thresholds of<br>maximal WT (99.7%PI)* | 4(1-10) | 99.5(98-99.9) | 0.52(0.47-0.56) | 60(20-90) | 83(82-83) | <0.0001 |
| WTSD | 64(53-75) | 99(97-99.7) | 0.82(0.78-0.85) | 93(83-97) | 93(91-95) | - |
| ED minimal WT | 1(0-7) | 97(94-98) | 0.49(0.44-0.54) | 7(1-36) | 82(81-83) | <0.0001 |
| Maximal-minimal WT | 56(44-67) | 90(86-92) | 0.72(0.68-0.77) | 53(45-62) | 90(88-92) | 0.0006 |
| ED mean WT | 5(1-12) | 97(94-98) | 0.51(0.46-0.55) | 82(82-83) | 83(81-84) | 0.0001 |
| ED maximal WT index | 18(10-28) | 94.5(91-96) | 0.56(0.52-0.61) | 40(27-56) | 84(83-86) | 0.0001 |
| Maximal apex/base ratio | 17(10-27) | 100(99-100) | 0.59(0.54-0.63) | 100(77-100) | 85(83-86) | <0.0001 |

\* Based on Shiwani et al (ref 9). ED, end-diastolic, WT wall thickness, SD standard deviation. With the exception of demographic-based personalized threshold, the cut-off of other parameters are shown in table 4.
ED maximal WT not included because it was used for diagnosis of all patients with HCM ( $\geq 15$ mm or $\geq 13$ mm in those with family history of HCM or with sarcomere pathogen mutation) and it was $< 13$ mm in all the mutation carriers without a diagnosis of HCM.

All patients with an HCM diagnosis based on current guidelines exhibited, for definition, either a maximal ED WT ≥13 mm (if carriers of pathogenic sarcomeric mutations) or ≥15 mm. Then, maximal ED WT was evaluated only indexed by BSA.

#### As shown in the same table, the Demographic-Based Personalized Threshold with a cut-off

>95th percentile demonstrated a sensitivity of 84% (79–87) and a specificity of 99.4% (98–99), with an AUC of 0.91 (0.89–0.94) for distinguishing HCM patients from healthy controls. This parameters identified 248 out of 297 HCM with 2 false positive. Compared to that parameter, the maximal ED WT indexed by BSA had a greater sensitivity (97%, CI 95-99), a lower specifity (89.5%, CI 86-92), and a non significantly different AUC (0.93, CI 0.91-0.95, p = 0.22). The WTSD showed a high sensitivity (97%, CI 94–98) and specificity (99%, CI 97–99.7), with an AUC of 0.98(0.96-0.99) that was significantly greater than that of all other parameters.

WTSD allowed identification of 287 out of 297 HCM patients with 4 false positive.

### Wall-thickness derived parameters in the whole population of mutation carriers without diagnosis of HCM

The Demographic-Based Personalized Threshold with a >95th percentile cut-off confirmed an excellent specificity (99.4%, CI 98–99), but with a very low sensitivity (11%, CI 5–20) and an AUC of 0.55 (0.51–0.60) to distinguish carriers of pathogenic sarcomeric mutations without an HCM diagnosis from healthy controls, allowing identification of only 9 out of 82 mutation carriers and with 2 false-positive cases. WTSD proved to be the most effective parameter also in this analysis, showing a sensitivity of 64% (53–75), a specificity of 99% (97–99.7), and an AUC of 0.82 (0.78–0.85), which was significantly superior to all other parameters. Indeed, WTSDallowed identification of 53 out of 82 mutation carriers, with only 4 false-positive cases. **Figure 2** shows the box-and-whisker plot illustrating differences in WTSD between mutation carriers and healthy controls across different age groups. WTSD was significantly higher in mutation carriers than in healthy controls for each class of age. An apex/base ratio >1 was identified in 14 mutation carriers (17%), but in none of the patients did the ED wall thickness of the apical segments indexed to BSA exceed the reference cut-off values proposed by Hughes and colleagues (10). Myocardial crypts were found in 22 (27%) and myocardial bridge of left anterior descending artery in 15 (18%) mutation carriers.

**Figure 2:**
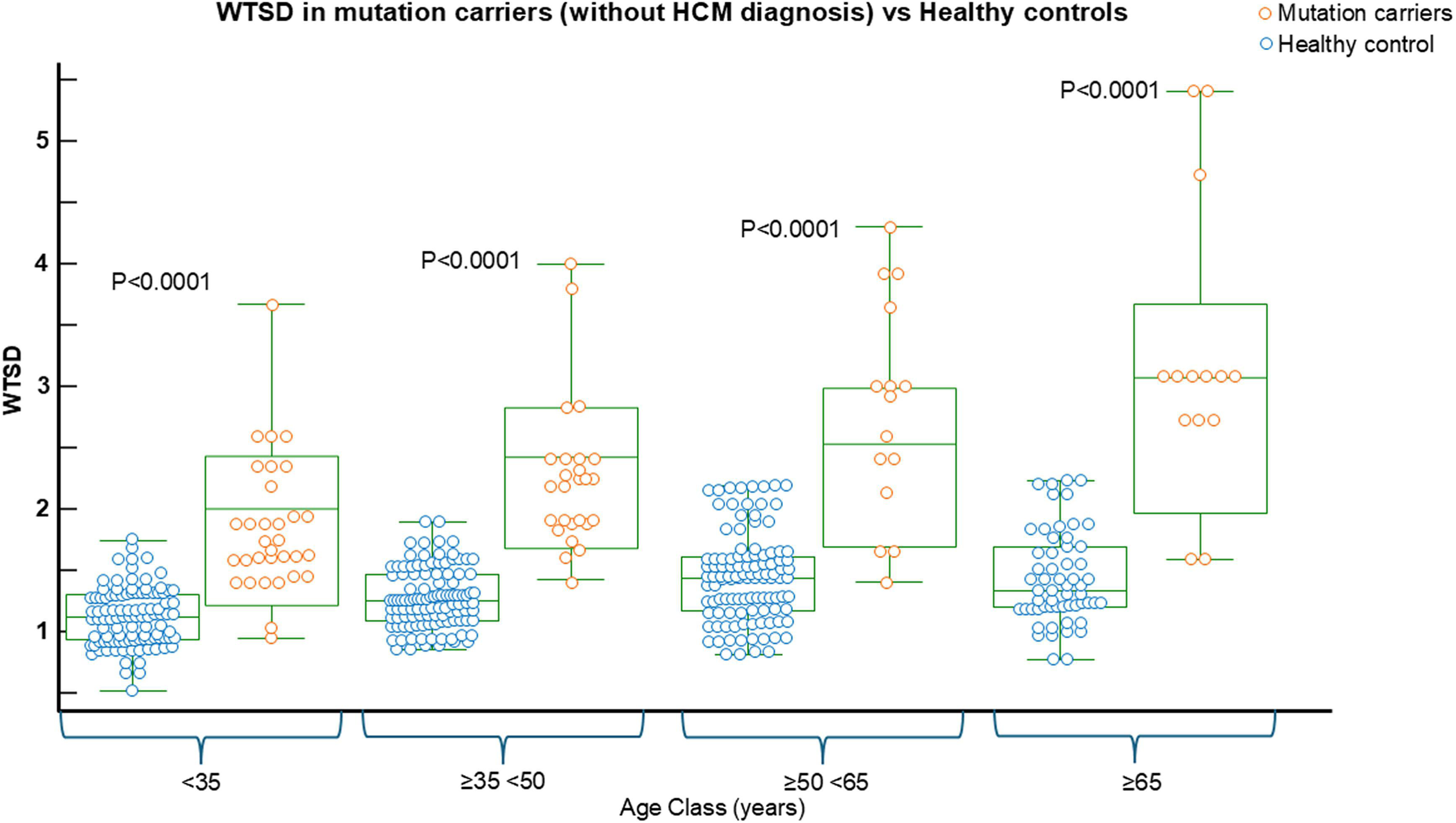
Box-and-whisker plot comparing WTSD between carriers of pathogenic sarcomeric mutations (without a diagnosis of HCM) and healthy controls in different age groups. This graph, which includes data from both males and females, clearly shows that WTSD was significantly higher in mutation carriers than in controls across all age groups

### Wall-thickness derived parameters in the female population

In the female population of patients diagnosed with HCM (Table 5), the Demographic-Based Personalized Threshold with a >95th percentile cut-off and the ED maximal WT m indexed by BSA showed identical performance, identifying 85 out of 92 patients with only one false positive, achieving good sensitivity (92%, CI 84–97) and excellent specificity (99%, CI 94–99), with an AUC of 0.96 (CI 0.93–0.98) in both cases.

**Table 5:** Diagnostic accuracy of wall thickness derived parameters in females.

| **Females with a guideline-based diagnosis of HCM vs controls:** |  |  |  |  |  |  |
| --- | --- | --- | --- | --- | --- | --- |
|  | **Sensitivity** | **Specificity** | **AUC** | **PPV** | **NPV** | **AUC comparison vs WTSD (P value)** |
| Demographic-Based Personalized Thresholds of maximal WT (95%PI)* | 92(84-97) | 99(97-99.9) | 0.96(0.93-0.98) | 98.8(92-99.8) | 96(92-99) | 0.012 |
| Demographic-Based Personalized Thresholds of maximal WT (99.7%PI)* | 81(72-89) | 99(95-99.9) | 0.90(0.86-0.94) | 98.6(91-99.8) | 90.7(86-94) | <0.0001 |
| WTSD | 98.9(94-99.9) | 100(98-100) | 0.995(0.98-1.0) | 100(96-100) | 99(96-99.9) | - |
| ED minimal WT | 7(1-14) | 97(93-99) | 0.52(0.46-0.58) | 54(27-79) | 65(63-66) | <0.0001 |
| Maximal-minimal WT | 98.9(94-99.9) | 90(85-94) | 0.95(0.91-0.97) | 85(78-90) | 99(95-99.9) | 0.005 |
| ED mean WT | 46(35-56) | 96(92-98.6) | 0.71(0.65-0.77) | 88(76-94) | 76(72-79) | <0.0001 |
| ED maximal WT indexed by BSA | 92(84-97) | 99(97-99.9) | 0.96(0.93-0.98) | 98.8(92-99.8) | 96(92-98) | <0.0001 |
| **Female carriers of sarcomere pathogen mutation without a HCM diagnosis vs controls:** |  |  |  |  |  |  |
|  | **Sensitivity** | **Specificity** | **AUC** | **PPV** | **NPV** | **AUC comparison vs WTSD (P value)** |
| Demographic-Based Personalized Thresholds of maximal WT (95%PI)* | 7(2-18) | 98(96-99) | 0.53(0.46-0.60) | 67(27-91) | 77(75-78) | <0.0001 |
| Demographic-Based Personalized Thresholds of maximal WT (99.7%PI)* | 2(1-10) | 99(95-99.9) | 0.50(0.43-0.57) | 33(4-84) | 76(75-76) | <0.0001 |
| WTSD | 74(60-85) | 99(97-99.9) | 0.87(0.82-0.91) | 98(85-99.6) | 92(85-99.6) | - |
| ED minimal WT | 0 | 95(91-98) | 0.47(0.41-0.54) | 0 | 75(73-75) | <0.0001 |
| Maximal-minimal WT | 48(34-62) | 93(88-96) | 0.71(0.64-0.76) | 68(54-80) | 84(79-88) | <0.0001 |
| ED mean WT | 6(1-15) | 99(98-99.8) | 0.53(0.49-0.55) | 33(11-66) | 96(95-96) | <0.0001 |
| ED maximal WT indexed by BSA | 11(4-22) | 98(94-99.6) | 0.55(0.48-0.61) | 67(34-88) | 77(75-79) | <0.0001 |

\* Based on Shiwani et al (ref 9). ED, end-diastolic, WT wall thickness. With the exception of demographic-based personalized threshold, the cut-off of other parameters are shown in table 4. ED maximal WT not included because it was used for diagnosis of all patients with HCM ( $\geq 15$ mm or $\geq 13$ mm in those with family history of HCM or with sarcomere pathogen mutation) and it was $< 13$ mm in all the mutation carriers without a diagnosis of HCM.

WTSD nevertheless proved to be the best parameter, identifying 91 out of 92 patients without any false positives, achieving a sensitivity of 98.9% (CI 94–99.9), a specificity of 100% (CI 98–100), and an AUC of 0.995 (CI 0.98–1.0), which was significantly superior to all other parameters. **Figure 3** shows the box-and-whisker plot illustrating differences in WTSD between females with HCM and healthy controls across different age groups.

**Figure 3:**
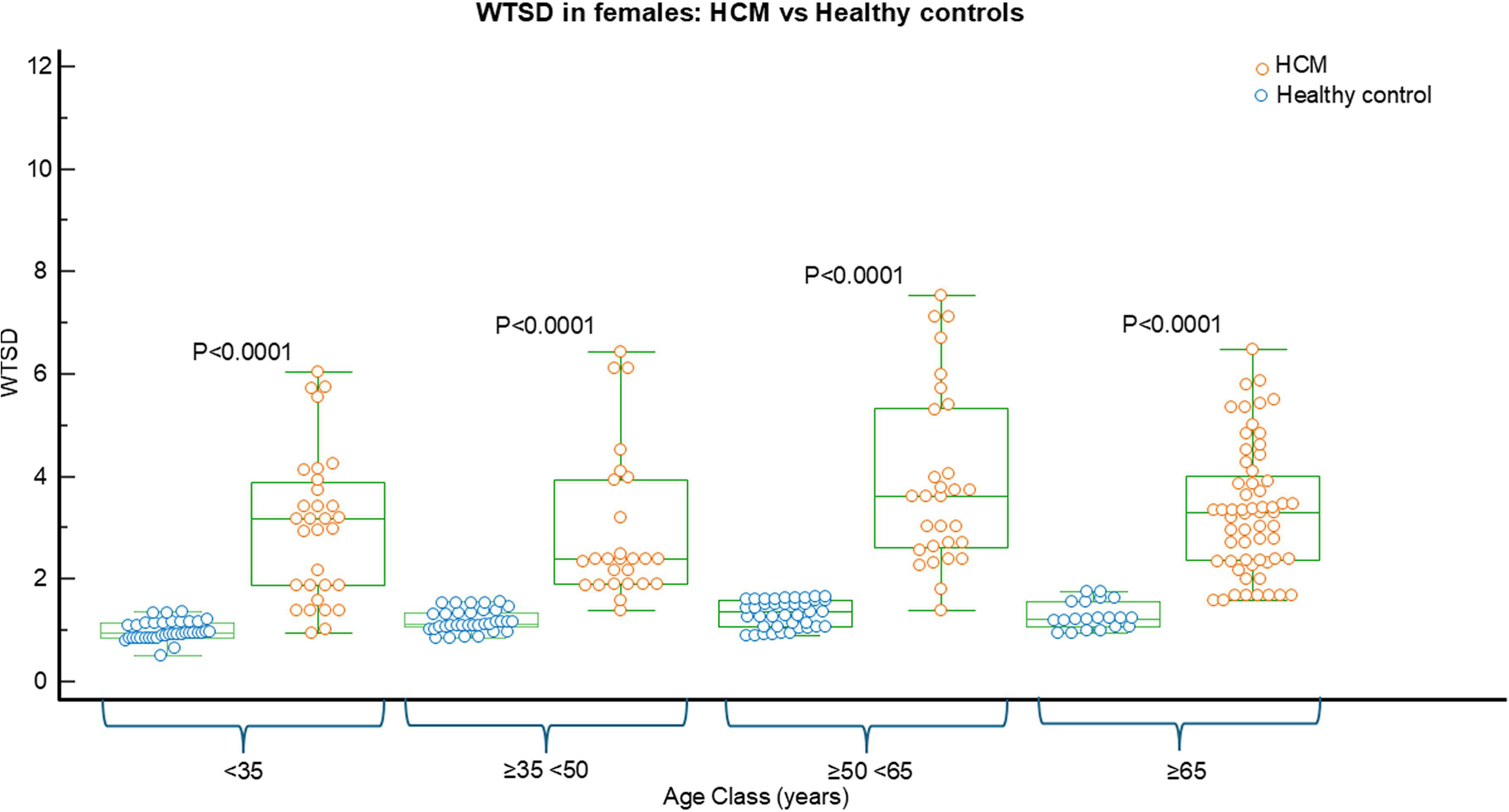
Box-and-whisker plot comparing WTSD between females patients with HCM and sex-matched healthy controls in different age groups. This graph clearly shows that WTSD was significantly higher in females with HCM than in controls across all age groups

The diagnostic effectiveness of WTSD was particularly evident in female carriers of pathogenic sarcomeric mutations without an HCM diagnosis (table 5), in whom this parameter identified 40 out of 54 patients with one false positive, achieving a sensitivity of 74% (CI 60–85), a specificity of 99% (CI 97–99.9), and an AUC of 0.87 (CI 0.82–0.91), which was significantly higher than in all other parameters (table 5). In fact, except for the end-diastolic maximal–minimal WT difference (AUC 0.71, CI 0.64-0.76) none of the other parameters reached statistical significance for the identification of mutation carriers compared to healthy controls.

### Wall-thickness derived parameters in the male population

In male subjects (Table 6) diagnosed with HCM, similar results observed in females were reproduced. In particular, the Demographic-Based Personalized Threshold with a >95th percentile cut-off and the maximal WT indexed by BSA demonstrated excellent specificity (100%, CI 98–100, and 95%, CI 94–98, respectively) and good sensitivity (81%, CI 75–87, and 88%, CI 83–92, respectively), with no significant difference in AUC (0.91, CI 0.88–0.93, and 0.92, CI 0.89–0.94, respectively; p = 0.59).

**Table 6:**
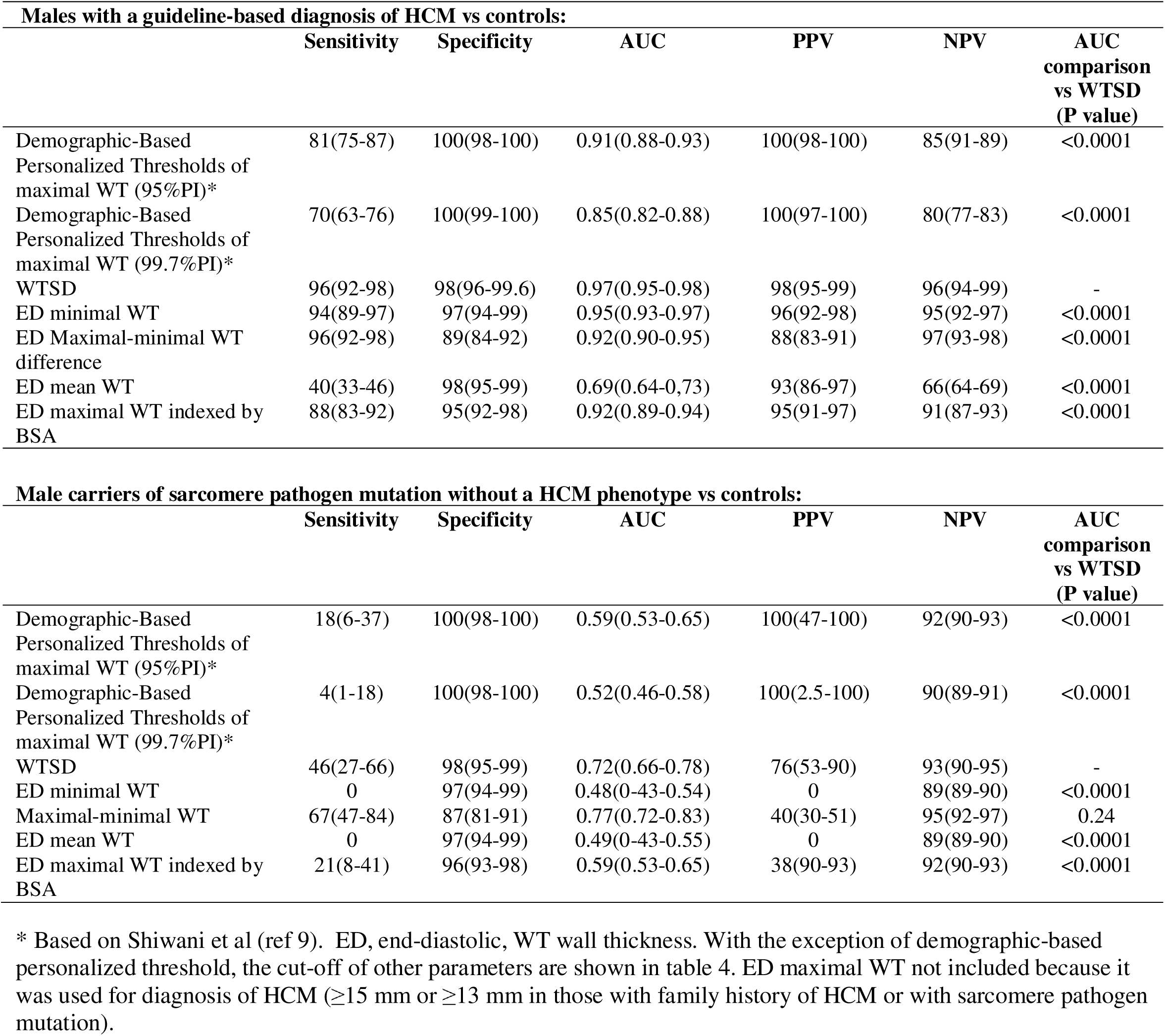
Diagnostic accuracy of wall thickness derived in males.

The Demographic-Based Personalized Threshold with a >95th percentile cut-off correctly identified 167 out of 205 male HCM patients without any false positives, while the maximal WT indexed by BSA identified 180 out of 205 patients but with 10 false positives.

Good diagnostic performance was also observed for the ED minimal WT- (AUC 0.95, CI 0.93– 0.97) and for the maximal-minimal ED WT difference (AUC 0.92, CI 0.90–0.95). However, in this group as well, WTSD proved to be the best-performing parameter, identifying 196 out of 205 patients with only 4 false positives, and showing an AUC of 0.97 (CI 0.95–0.98), which was significantly higher than that of all other parameters (table 6). **Figure 4** shows the box-and-whisker plot illustrating differences in WTSD between males with HCM and healthy controls across different age groups.

**Figure 4:**
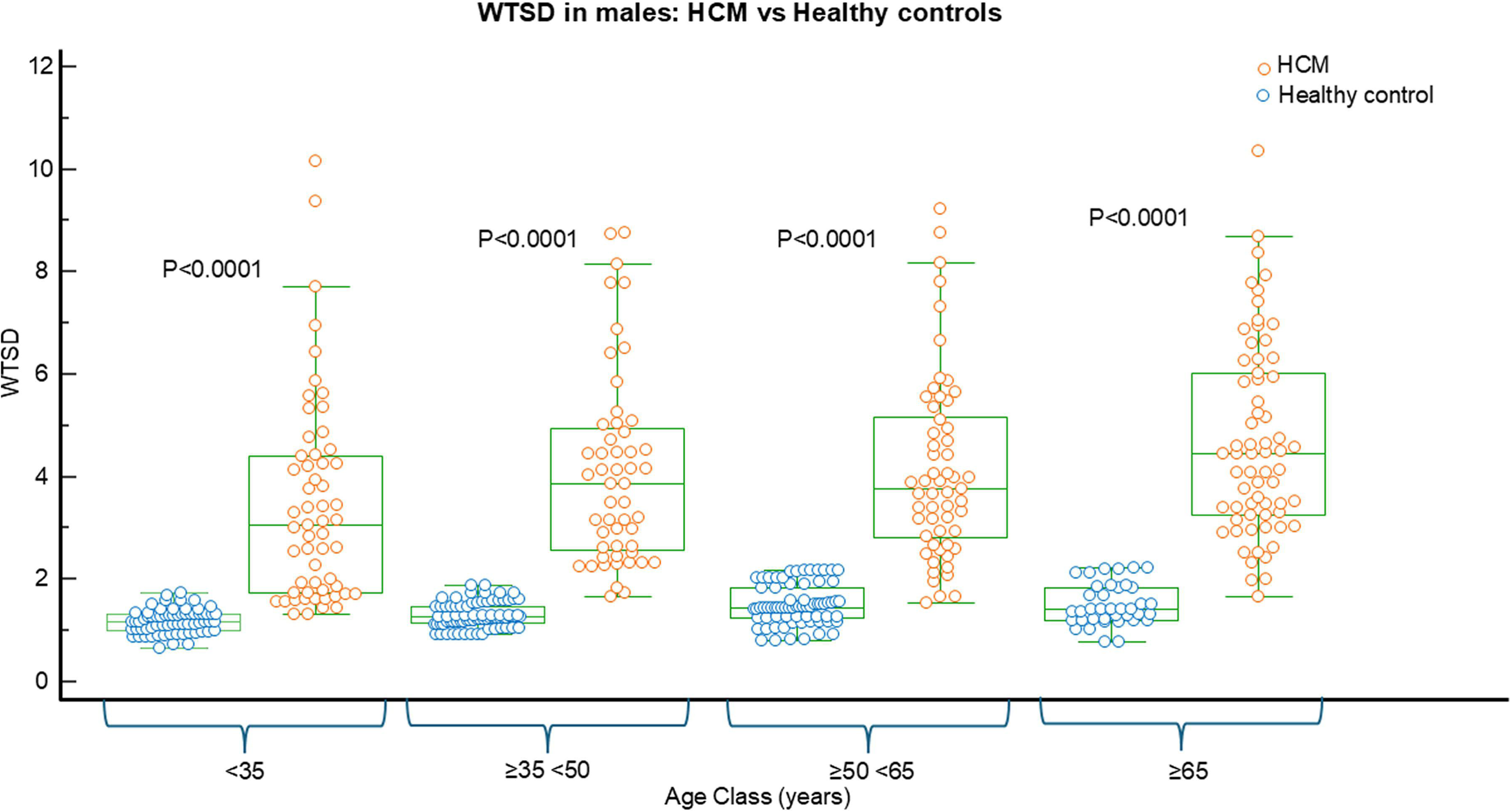
Box-and-whisker plot comparing WTSD between males patients with HCM and sex-matched healthy controls in different age groups. This graph clearly shows that WTSD was significantly higher in males with HCM than in controls across all age groups

WTSD and the maximal–minimal ED WT difference were the best parameters to identify male carriers of pathogenic sarcomeric mutations without an HCM diagnosis with respectively AUC of 0.72(CI 0.66-0.78) and 0.77(0.72-0.83), p = 0.23. WTSD allowed identification of 13 out of 28 male mutation carriers with 4 false positive, while the maximal–minimal ED WT difference identified 19 out of 28 carriers with 28 false positive.

### Wall-thickness derived parameters in whole population of patients with pathogen sarcomeric mutations

Overall, 246 patients had a pathogen sarcomeric mutation (including those with a definite diagnosis of HCM and mutation carriers). The effectiveness to identify subjects with pathogen sarcomeric mutations (including both those with definite diagnosis of HCM and mutation carriers without a diagnosis) of wall thickness derived parameters is shown in table 7. WTSD was the parameter with best performance allowing to identify 212 out 246 patients with mutation with 4 false positive, achieving a sensitivity of 86% (CI 81-90), a specificity of 99% (CI 97-99.7) and AUC of 0.93(0.90- 0.95).

**Table 7:** Diagnostic accuracy of wall thickness- derived parameters in all patients with sarcomere pathogen mutation vs controls.

|  | Sensitivity | Specificity | AUC | PPV | NPV | AUC comparison vs WTSD (P value) |
| --- | --- | --- | --- | --- | --- | --- |
| Demographic-Based Personalized Thresholds of maximal WT (95%PI)* | 63(56-69) | 99(98-99.8) | 0.81(0.78-0.84) | 98(94-99) | 81(77-83) | <0.0001 |
| Demographic-Based Personalized Thresholds of maximal WT (99.7%PI)* | 59(52-65) | 99(98-99.8) | 0.79(0.76-0.82) | 98(94-99) | 79(76-81) | <0.0001 |
| WTSD | 86(81-90) | 99(97-99.7) | 0.93(0.90-0.95) | 98(95-99) | 92(89-94) | - |
| ED minimal WT | 4(2-7) | 97(94-98) | 0.50(0.46-0.54) | 41(23-61) | 61(60-62) | <0.0001 |
| Maximal-minimal WT | 83(79-87) | 89(86-92) | 0.86(0.84-0.89) | 83(79-87) | 89(86-92) | <0.0001 |
| ED mean WT | 35(29-42) | 97(96-98) | 0.66(0.62-0.70) | 88(80-93) | 69(68-72) | <0.0001 |
| ED maximal WT indexed by BSA | 67(60-73) | 95(92-97) | 0.81(0.77-0.84) | 89(79-84) | 82(79-84) | <0.0001 |

\* Based on Shiwani et al (ref 9). ED, end-diastolic, WT wall thickness. With the exception of demographic-based personalized threshold, the cut-off of other parameters are shown in table 4. ED maximal WT not included because it was used for diagnosis of HCM ( $\geq 15$ mm or $\geq 13$ mm in those with family history of HCM or with sarcomere pathogen mutation).

The **Central Illustration** shows three representative female cases (a healthy control, a mutation carrier, and a patient with overt HCM) together with the summary table reporting the diagnostic accuracy of WTSD across the different patient groups analyzed and in the overall population.

## DISCUSSION

The results of this study suggest that increased left ventricular wall thickness heterogeneity, as reflected by high WTSD values, represents a phenotypic hallmark of sarcomeric cardiomyopathy. WTSD was increased in patients with definite HCM compared to age- and sex-matched healthy controls, and it was also elevated in the majority (64%) of individuals carrying a pathogenic sarcomeric mutation but without left ventricular hypertrophy. In both scenarios, patients with definite HCM and individuals carrying a pathogenic mutation, an elevated WTSD suggests the presence of significant wall thickness heterogeneity. In patients with definite HCM, this heterogeneity is driven by marked differences in wall thickness among hypertrophic segments, as well as the coexistence of hypertrophic segments with segments of normal or even reduced thickness (although HCM patients did not show differences in minimal WT compared to healthy controls). In mutation carriers, on the other hand, no hypertrophic segments are present; rather, heterogeneity results from the coexistence of normal and thinned segments. Indeed, mutation carriers showed significantly lower minimal WT values compared to both healthy controls and HCM patients, while their maximal WT did not differ from that of healthy individuals.

The observation that HCM often exhibits asymmetric or regional hypertrophy with adjacent areas of relative thinning is not novel and reflects the typical pattern of hypertrophic development, which frequently follows a spiral trajectory from base to apex in a counterclockwise direction (14). In HCM, the LV mass index is often increased, but it may also be within the normal range due to the coexistence of wall hypertrophy and areas of thinning (15)

In post-mortem CMR,hearts of patients with HCM and pathogen sarcomeric mutation have significantly greater wall thickness dispersion than both normal hearts and non-HCM hypertrophy controls (12). Similarly, a living CMR study of genotype-positive relatives reported subtle structural areas of abnormalities in all carriers, including abrupt regional changes in wall thickness (>50% difference between adjacent segments in 39% of cases) and focal “inappropriate” thinning in 50%, features that is never observed in healthy controls (16). Such heterogeneity in wall architecture appears to be a distinctive morphologic signature of HCM, evident even when absolute wall thickness remains within normal limits.

These results also highlight the limitations of conventional diagnostic metrics in genotype-positive individuals. Current guidelines rely on an end-diastolic wall thickness ≥15 mm (or ≥13 mm with a familial mutation) to define HCM (2), a threshold that by definition misses those in the preclinical phase. Many sarcomere mutation carriers show no hypertrophy for years, with one longitudinal study estimating only 46% penetrance at 15-year follow-up (17). Notably, a subset of 32% of carriers in that study fulfilled HCM criteria by CMR (often due to apical or subtle hypertrophy) that were not apparent by echocardiography, emphasizing the superior sensitivity of CMR in early disease detection. As previous reported, recent efforts to improve diagnostic sensitivity have proposed adjusting wall thickness cut-offs for demographic factors or indexing hypertrophy to body size. In the previously mentioned study, Shiwani et al created a personalized threshold accounting for sex, age, and BSA, showing to maintain high specificity(9). However, even such refined metrics remain predicated on surpassing a hypertrophy threshold. In our cohort, the demographic-based 95th-percentile cutoff identified only 11% of mutation carriers, whereas WTSD—was increased in64% of them, achieving an maintaining an excellent diagnostic specificity.

Genotype-positive individuals traditionally classified as “phenotype-negative” may, in fact, represent an early phenotypic stage of HCM that eludes current diagnostic thresholds. Our findings, together with emerging imaging data, challenge the binary distinction between genotype-positive/phenotype-negative and overt HCM, suggesting instead a continuum of disease expression. A substantial proportion of mutation carriers already exhibit measurable structural alterations, an increased wall thickness heterogeneity, despite not meeting the absolute wall thickness cutoffs of 13–15 mm. The WTSD provides a sensitive and reproducible means to detect these subtle yet clinically relevant abnormalities. The ability to identify mutation carriers who have not yet developed the hypertrophic phenotype may have important prognostic implications. It is reasonable to assume that some of these individuals, particularly the younger ones, may go on to develop hypertrophy over time. However, we cannot determine if or how many mutation carriers would eventually develop the phenotype, as the design of the present study did not include longitudinal follow-up of these patients. Future studies are needed to assess whether the novel therapies for HCM, as cardiac myosin inhibitors, can prevent the development of the hypertrophic phenotype and reduce the associated risks in mutation carriers with marked wall thickness heterogeneity. The implications are also significant for individuals who may never develop the phenotype, as the imaging-based identification of a possible sarcomeric mutation would prompt genetic testing in first-degree relatives, allowing for the early identification of those who may eventually develop the disease.

Sex-specific results of the present study provided additional insight into relevant diagnostic implications. Female HCM patients are known to present later and with more advanced disease than males (7,8), an observation often attributed to smaller baseline LV size and under-detection by WT criteria. Our results reflect this disparity: by applying sex-specific WTSD thresholds, we achieved near-perfect sensitivity and specificity in women with overt HCM and, importantly, identified approximately three-quarters of female mutation carriers who would otherwise have remained undiagnosed. In contrast, traditional cut-offs (even when personalized) tend to miss women until significant hypertrophy has developed. These findings align with prior reports that women with HCM have been historically under-recognized until a later stage (8). By comparison, male mutation carriers posed a greater challenge. WTSD still outperformed other metrics in men but picked up a lower proportion of male carriers than female carriers. One explanation is that male hearts have higher absolute wall thickness norms and often develop HCM phenotype earlier in life; 17) thus, fewer men remain in a subtle pre-hypertrophic state, and those who do may have more homogeneous wall thickness. In fact, male sex has been associated with a nearly threefold higher hazard of transitioning from carrier status to overt HCM (17), and not surprisingly, only about one-third of the mutation carriers in our study were male. . This suggests that while heterogeneity-based diagnostics improve detection in both sexes, they are especially impactful for females, who experience slower, less diffuse hypertrophy progression. Tailoring diagnostic approaches to sex is crucial,as evidenced by Shiwani et al.’s personalized thresholds (9), and WTSD provides a promising sex-sensitive marker that narrows the diagnostic gap between men and women.

An intriguing aspect of our cohort of mutation carriers was the presence of abnormally thin myocardial segments alongside mildly thickened ones, contributing to the elevated WTSD. Such localized wall thinning has been noted in preclinical HCM, who reported that genotype-positive individuals frequently exhibit discrete basal septal crypts or clefts. Maron et al. (18) found myocardial crypts on CMR in 61% of genotype-positive/phenotype-negative subjects (versus only 4% of overt HCM patients and none of controls), underscoring that these microstructural abnormalities are a hallmark of the prephenotypic state of this cardiomyopathy. Crypts are typically confined to the basal ventricular septum or posterior wall and are not associated with fibrosis or systolic dysfunction, suggesting they represent benign developmental aberrations or early remodeling. Nevertheless, their strong enrichment in mutation carriers highlights how the sarcomere mutation can subtly alter cardiac morphology long before gross hypertrophy. Wall thinning may coexists with crypts but also with intracavitary hypertrabeculation. In the present study, we measured the WT by considering only the “compact” myocardium and excluding such trabeculations. Wall thinning, crypts, and intracavitary hypertrabeculation may represent signs of regionally ’underdeveloped’ myocardium accompanying incipient hypertrophic changes. This may be reflected by significantly lower minimal and mean WT values in mutation carriers compared to healthy controls. It is conceivable that such thinning reflects uneven hypertrophy. Regardless of the mechanism, the coexistence of segmental hypertrophy and thinning is a unique structural fingerprint of HCM. Clinically, this reinforces the idea that **any** significant heterogeneity in wall thickness,even if absolute values are normal,should raise suspicion in at-risk patients. In practice, CMR is invaluable for detecting these subtle markers (crypts, bridges, wall thickness abrupt shifts) that echocardiography may overlook. Advanced echocardiographic techniques have also shown promise in this arena: for instance, abnormal diastolic flow propagation and heightened apical rotation can distinguish mutation carriers with 83% sensitivity before hypertrophy develops. (19). Future studies are needed to assess the combined diagnostic role to identify mutation carriers of these novel functional echocardiographic techniques with the evaluation of wall thickness heterogeneity through WTSD measured by CMR.

Some study limitations need to be mentioned. First, the results of the present study were obtained in relatively small patient cohorts, particularly considering the limited number of mutation carriers. Larger population-based studies are needed to confirm our findings in both mutation carriers and patients with overt HCM. Second, this study compared healthy controls with mutation carriers; therefore, the diagnostic performance of WTSD and other parameters should be tested in unselected patient populations to evaluate their ability to identify mutation carriers in real-world clinical settings. Third, our study did not assess the role of WT-derived parameters in other cardiomyopathies with a hypertrophic phenotype, such as cardiac amyloidosis, Fabry disease, glycogen storage diseases, or hypertensive heart disease. Further studies are needed to investigate this aspect. Nonetheless, even if WTSD were found to be altered in these other cardiac conditions and thus unable to distinguish sarcomeric disease from other etiologies, this would not undermine its ability to differentiate healthy individuals from those with pathological myocardial involvement.Finally, although we did not exclude healthy controls who engaged in regular recreational physical activity, we cannot confirm or rule out whether WTSD can distinguish between HCM and elite athletes with physiological (eccentric) hypertrophy. Further studies are needed to clarify this aspect as well.

## CONCLUSIONS

Our study demonstrates that wall thickness heterogeneity, quantified as WTSD, is a powerful and versatile imaging biomarker across the full spectrum of sarcomeric HCM. WTSD outperforms conventional wall thickness–based criteria not only in detecting early phenotypic expression among genotype-positive individuals, but also in accurately identifying overt disease. These findings, if confirmed by future large population study, may support the integration of WTSD into diagnostic algorithms, promoting a shift from rigid threshold-based definitions toward a more dynamic, sex-based, and sensitive approach to HCM diagnosis and risk stratification.

## Data Availability

All data produced in the present study are available upon reasonable request to the authors

## Aknowledge

all authors meet the authorship criteria

---

**Central Illustration:**
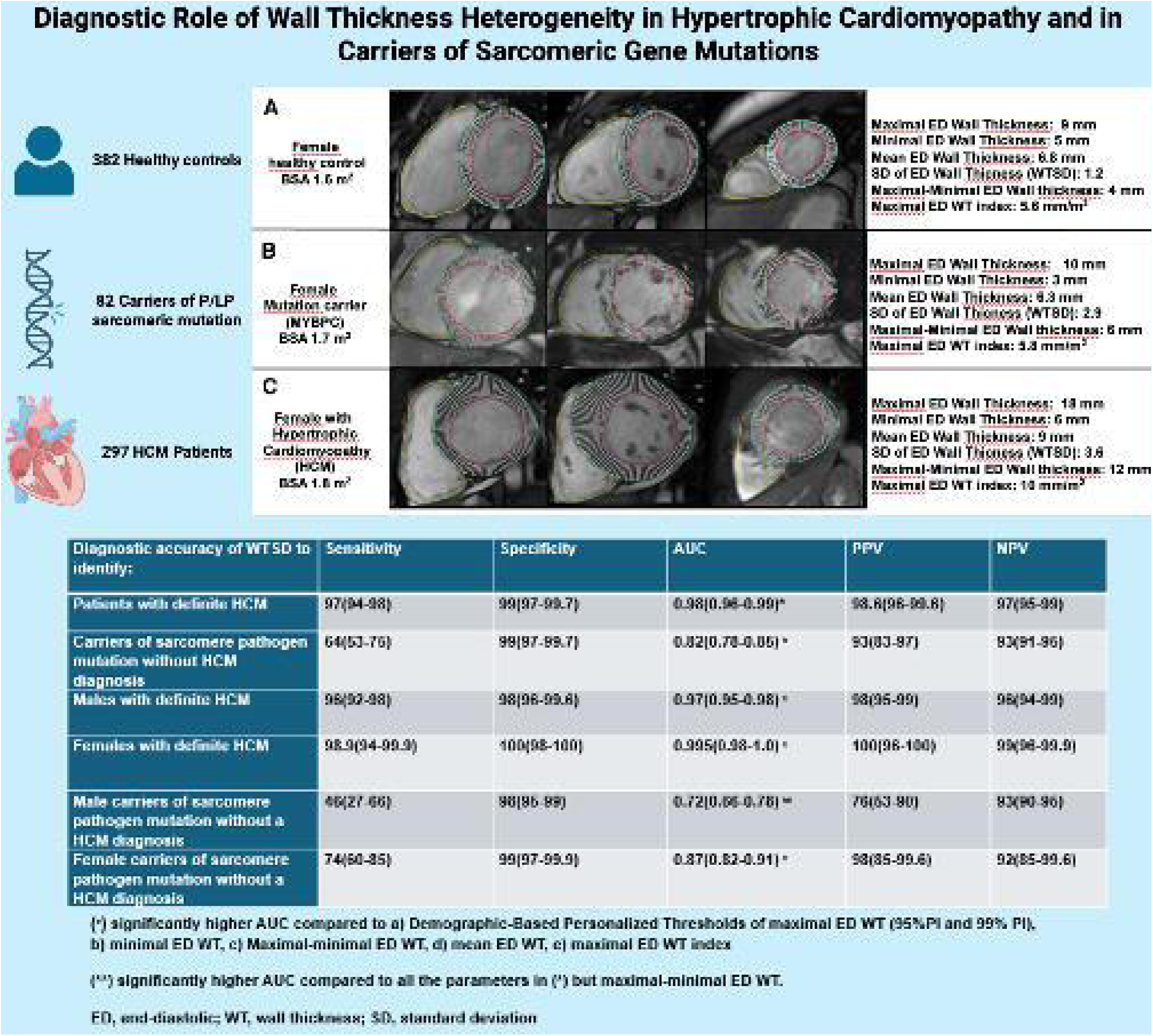
In the upper part examples of image analysis in a healthy control, a mutation carrier, and a patient with HCM are reported. A panel shows three short-axis images from a female healthy control aged 36–40 years with a body surface area (BSA) of 1.6 m². B panel displays images from a female mutation carrier aged 36–40 years (pathogenic MYBPC3 mutation) with a BSA of 1.7 m². C panel shows images from a female with a definitive diagnosis of HCM aged 26–30 years. The rightmost panels display the corresponding wall thickness parameters for each case. As shown, the maximal end-diastolic (ED) wall thickness (WT) was within the normal range in both the healthy control and the mutation carrier, even when considering Demographic-Adjusted Left Ventricular Hypertrophy Thresholds (ref. 9). The mutation carrier exhibited a lower minimal WT and a greater maximal–minimal WT difference compared to the control. Notably, the standard deviation of wall thickness (WTSD) was more than twice as high in the mutation carrier than in the healthy control. **In the lower part the summary table reporting the diagnostic accuracy of WTSD across the different patient groups analyzed and in the overall population is reported**

